# c-Triadem: A constrained, explainable deep learning model to identify novel biomarkers in Alzheimer’s disease

**DOI:** 10.1101/2024.11.19.24317595

**Authors:** Sherlyn Jemimah, Ferial Abuhantash, Aamna AlShehhi

## Abstract

Alzheimer’s disease (AD) is a neurodegenerative disorder that requires early diagnosis for effective management. However, issues with currently available diagnostic biomarkers preclude early diagnosis, necessitating the development of alternative biomarkers and methods, such as blood-based diagnostics. We propose c-Triadem (constrained triple-input Alzheimer’s disease model), a novel deep neural network to identify potential blood-based biomarkers for AD and predict mild cognitive impairment (MCI) and AD with high accuracy. The model utilizes genotyping data, gene expression data, and clinical information to predict the disease status of participants, i.e., cognitively normal (CN), MCI, or AD. The nodes of the neural network represent genes and their related pathways, and the edges represent known relationships among the genes and pathways. We trained the model with blood genotyping data, microarray, and clinical features from the Alzheimer’s Neuroimaging Disease Initiative (ADNI). We demonstrate that our model’s performance is superior to previous models with an AUC of 97% and accuracy of 89%. We then identified the most influential genes and clinical features for prediction using SHapley Additive exPlanations (SHAP). Our SHAP analysis shows that CASP9, LCK, and SDC3 SNPs and PINK1, ATG5, and ubiquitin (UBB, UBC) expression have a higher impact on model performance. Our model has facilitated the identification of potential blood-based genetic markers of DNA damage response and mitophagy in affected regions of the brain. The model can be used for detection and biomarker identification in other related dementias.

**Author Summary:** C-Triadem, our novel developed deep neural network, accurately predicts moderate cognitive impairment (MCI) and Alzheimer’s disease (AD) while identifying potential blood biomarkers for AD. Current diagnostic methods have limitations, emphasizing the critical need for early AD detection. Our model integrates genetic, gene expression, and clinical data to differentiate among cognitively normal individuals, MCI, and AD cases. Training and validation using Alzheimer’s Disease Neuroimaging Initiative (ADNI) data demonstrate superior performance, with a 97% AUC and 89% accuracy, surpassing previous models. SHapley Additive exPlanations (SHAP) analysis highlights key clinical features (e.g., MMSE scores, brain volume) and genes (e.g., CASP9, LCK, SDC3), revealing potential genetic markers and pathways in blood associated with AD. By incorporating Reactome pathways, our approach enhances interpretability, providing insight into the biological context of predictions. In summary, c-Triadem represents a significant advancement in AD diagnostics, enabling earlier and more accurate diagnoses for improved treatment strategies.

## Introduction

Alzheimer’s disease (AD) is the most common form of dementia, characterized by a gradual loss of cognition and memory. It is expected to affect around 78 million elderly by 2030. [1] While the APOE *ϵ*4 allele, mutations in presenilin-I and APP (amyloid precursor protein) are established genetic markers, [2] AD is considered a multifactorial, complex disease driven by both genetic and environmental factors. [3] The AD hallmarks include amyloid-*β* (A*β*) deposition and formation of neurofibrillary tangles (NFT) such as tau protein aggregates. [4] Its early symptoms are traced by the higher rates of neurodegeneration in the entorhinal cortex and hippocampus cornu ammonis 1 region, [5, 6] which correlated with increased NFT in this regions. [7]

AD hallmarks are used by the National Institute on Aging and Alzheimer’s Association (NIA-AA) to categorize existing AD biomarkers under A*β* deposition, pathologic tau, and neurodegeneration [AT(N)] research framework, enabling a biological definition of AD. [8] However, the diagnostic modalities for AT(N) biomarkers (such as MRI (magnetic resonance imaging), PET (positron emission tomography), and lumbar puncture) are known to be expensive or invasive. Padala and Newhouse [9] -knowing that MRI and PET scans can cost up to US$ 8,000 and US$ 3,400 respectively. In contrast, blood tests are relatively cost-effective (up to US$ 1,250). Therefore, clinical diagnosis through blood sampling has been proposed to make testing more accessible and affordable for the patient population, and enable routine monitoring [9]; as well as recent research has focussed on elucidating blood-based biomarkers, to facilitate less-invasive diagnostic tests for AD. [10–13]

Nonetheless, several hurdles remain for conventional biomarker testing in blood samples. One major issue is the quantitation of extremely low levels of A*β* and tau proteins against a background with high levels of plasma proteins, such as albumin and immunoglobulin. The low levels of A*β* and tau may be further subject to metabolization and clearing by physiological processes. [14] While the issue may be overcome with ultrasensitive assays, [15] the considerable degree of overlap in the levels of A*β* between AD patients and cognitively normal individuals precludes its use as a definitive blood marker by itself. [16] Moreover, existing biomarkers for diagnosis require significant levels of A*β* deposition and/or tau pathology for detection, precluding early diagnosis which is imperative for effective treatment. [17]

The disadvantages of conventional biomarkers in blood-based diagnostics highlight the need to develop novel methods and biomarkers which can facilitate accurate, early diagnosis, based on experimental evidence of changes in the blood cells of patients with MCI (mild cognitive impairment, ie., prodromal stage) and advanced AD. [18] Polygenic risk scores capture the association of several gene loci with plasma biomarkers [19] and are shown to improve accuracy in distinguishing cognitively normal (CN) and AD cases. [20] Reddy et al. [21] showed that plasma mRNA levels of an eQTL (expression quantitative trait locus)-curated gene panel significantly increased diagnostic accuracy and may also have utility in discriminating between different types of dementia. Blood-based panels of potential biomarker genes have been identified using machine learning (ML) for early detection of AD. [17, 22]

With ML and artificial intelligence (AI), accurate and early diagnosis using multimodal, blood-based data is a distinct possibility. Several ML models have been proposed to distinguish between healthy CN, MCI, and advanced AD patients. Stamate et al. [23] utilized plasma metabolites in an XGBoost model to achieve an AUC of 0.89 in distinguishing between AD and CN cases. Logistic regression using plasma levels of inflammatory proteins enabled the differentiation of AD from controls (AUC 0.79) and MCI subjects (AUC 0.74). [24] A Random forest model of serum protein multiplex biomarker data attained an AUC of 0.91 in predicting AD and CN cases. [25] Qui et al. [26] proposed a deep learning framework that uses neuroimaging data and patient demographic information for binary classification of AD and cognitively normal participants with accuracy comparable to human experts. Advances in deep learning such as the development of the SHapley Additive exPlanations (SHAP) [27] algorithm enable the investigation of multi-omic data with transparent, biologically relevant models, which is important to engender trust and facilitate adoption by clinicians. [28]

In this study, we introduce c-Triadem, a constrained deep neural network multiclass-classifier, which incorporates prior biological information in the form of Reactome pathways to accurately classify samples as CN, MCI, or AD. Genotyping, gene expression, and clinical data from the Alzheimer’s Disease Neuroimaging Initiative (ADNI) [29] are used for training, validation, and testing. The significance of genetic and clinical features in the input is assessed by employing SHapley Additive exPlanations (SHAP). Further, we interrogate the intermediate layers to better understand model behavior and gain mechanistic insights into the role of the genes identified as potential biomarkers. The proposed methodology, experimental data, results, and conclusions are presented in the following sections.

## Results

### Clinical cohort characteristics

The cohort of selected ADNI participants includes 212 CN, 317 MCI, and 97 AD subjects. The clinical features after imputation are summarized in Tables 1 and 2. In general, the CN, MCI, and AD groups differ significantly in terms of mean age (p < 0.001) and proportion of female participants (p = 0.012). We also observe a higher proportion of AD patients with the APOE *ϵ*4 allele compared to MCI and CN subjects (p < 0.001). As expected, AD patients show significantly worse performance in neuropsychological tests (p < 0.001). Furthermore, measures of brain volume are lower in AD patients (p < 0.001) with the exception of ICV, which remains similar across the groups. Brain functioning measures are also significantly impacted in AD with higher levels of AV45 and CSF tau protein, along with lower FDG uptake (p < 0.001).

**Table 1.** ADNI demographics and brain test variable clinical data summary showing average values with standard deviation. Statistical significance (95% significance level) was tested using analysis of variance (ANOVA) for the age of onset and years of education, Chi-square contingency for gender proportions and APOE *ϵ*4 allele, and Kruskal-Wallis test for all other variables.

|  | CN | MCI | AD | P-value |
| --- | --- | --- | --- | --- |
| Number of samples | 212 | 317 | 97 |  |
| APOE ε4 allele present | 58 | 154 | 77 | <0.001 |
| Gender (female) | 105 | 131 | 31 | 0.012 |
| Age | 74.620±5.444 | 72.069±7.462 | 74.723±7.594 | <0.001 |
| Years of education | 16.231±2.675 | 15.911±2.747 | 16.175±2.912 | 0.384 |
| **Brain functioning and clinical tests** |  |  |  |  |
| FDG uptake | 1.299±0.112 | 1.267±0.128 | 1.045±0.150 | <0.001 |
| AV45 uptake | 1.103±0.182 | 1.193±0.226 | 1.341±0.243 | <0.001 |
| Level of tau protein in CSF | 247.913±81.605 | 266.968±122.224 | 374.673±143.266 | <0.001 |
| **Brain volume measurements from MRI** |  |  |  |  |
| Ventricles | 34517.162±18550.933 | 38657.239±22996.339 | 55538.765±25105.972 | <0.001 |
| Hippocampus | 7270.301±956.410 | 7031.596±1113.513 | 5436.083±1067.069 | <0.001 |
| Entorhinal complex | 3754.567±666.689 | 3681.1867±23.034 | 2610.140±694.886 | <0.001 |
| Fusiform complex | 18144.345±2461.023 | 18383.708±2776.218 | 15072.561±2330.017 | <0.001 |
| Mid-temporal | 19974.678±2681.839 | 20479.689±2777.653 | 16467.175±3162.854 | <0.001 |
| Intracranial volume (ICV) | 1500236.243±156407.973 | 1530645.222±151053.965 | 1546662.105±191022.847 | 0.0977 |
| Whole brain | 1018671.177±107104.069 | 1052311.884±109453.752 | 955524.111±118730.033 | <0.001 |

**Table 2.**
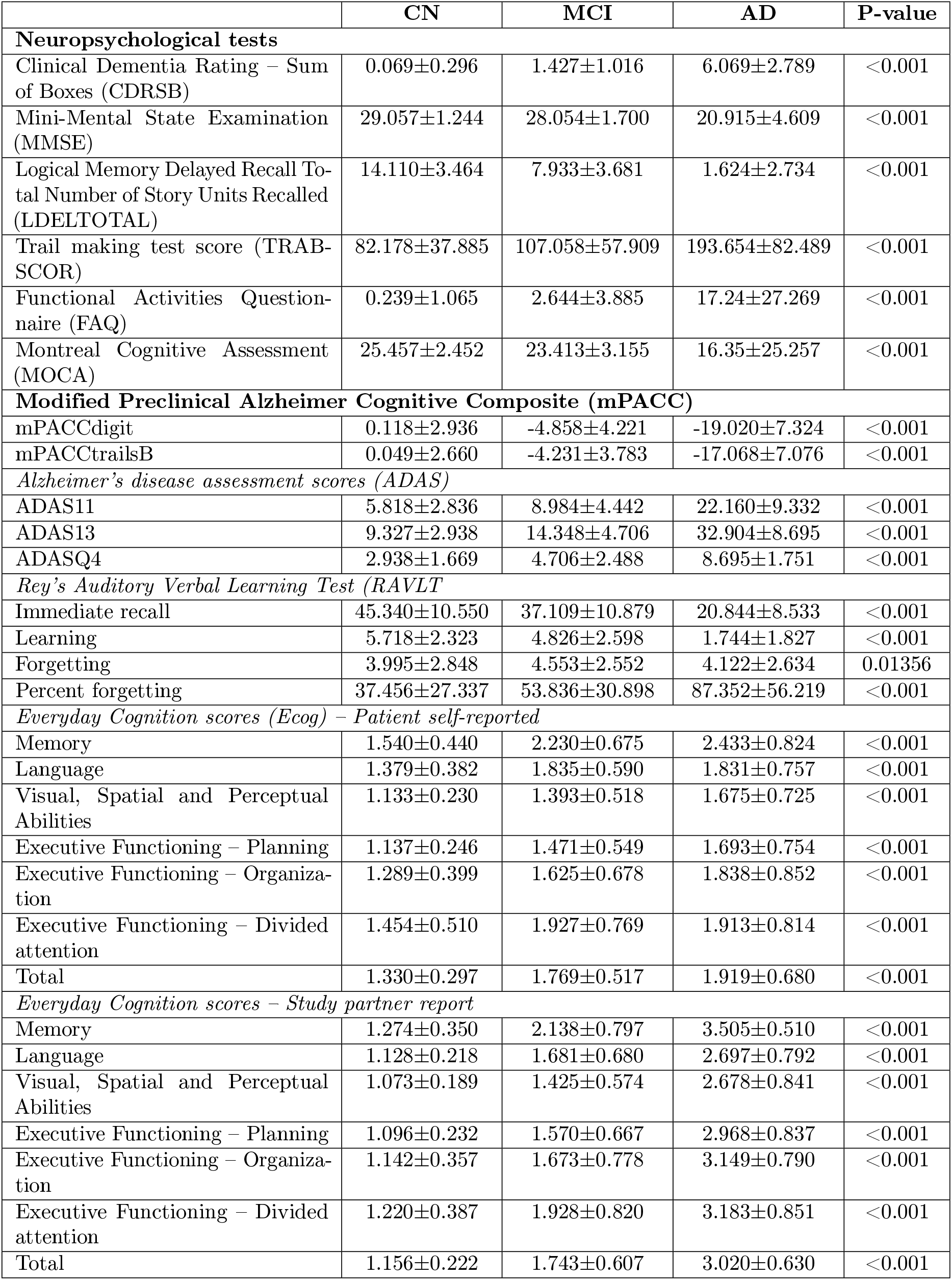
ADNI neuropsychological test clinical data summary showing average values with standard deviation. Statistical significance (95% significance level) was tested using analysis of Kruskal-Wallis test for all Neuropsychological test and other variables.

### Model performance

With c-Triadem, we achieved an accuracy of 89% and an AUC of 97% on the test data. We also developed an unconstrained dense network with similar architecture (accuracy 87% and AUC 96%) for comparison and were able to demonstrate c-Triadem’s superior performance. We also tested whether our model may be refined by additional pathway layers. Training our model with two additional pathway layers in each subnetwork produced comparable results with an AUC of 96% and an accuracy of 88%. The performance of our model compared to other available machine learning classifiers on ADNI data is shown in Table 3.

**Table 3.** Performance comparison of multiclass methods using ADNI data.

| Model | Inputs | Accuracy | AUC | F1 score | Reference |
| --- | --- | --- | --- | --- | --- |
| c-Triadem | 2=Clinical, SNPs and expression data | 0.893 | 0.972 | 0.893 | This work |
| Unconstrained dense network |  | 0.866 | 0.960 | 0.869 |  |
| Deep model | Clinical data only | 0.760 | NA | 0.760 | Venugopalan et al., 2021 [30] |
| Deep model | Clinical data + SNPs | 0.780 | NA | 0.780 | Venugopalan et al., 2021 [30] |
| Stage-wise deep network | SNPs only | 0.541 | NA | NA | Zhou et al., 2019 [31] |
| Gradient boosting classifier with Soft Impute | Clinical data only | 0.6503 | NA | 0.6305 | Aghili et al., 2022 [32] |

### Model explanation

To examine the importance of specific genes and clinical features in model prediction, we computed SHAP values in the constrained model for the three types of input, i.e., SNPs, gene expression, and clinical features. Beeswarm plots for the top 20 features in each type of input are provided in Figure 1.

**Fig 1.**
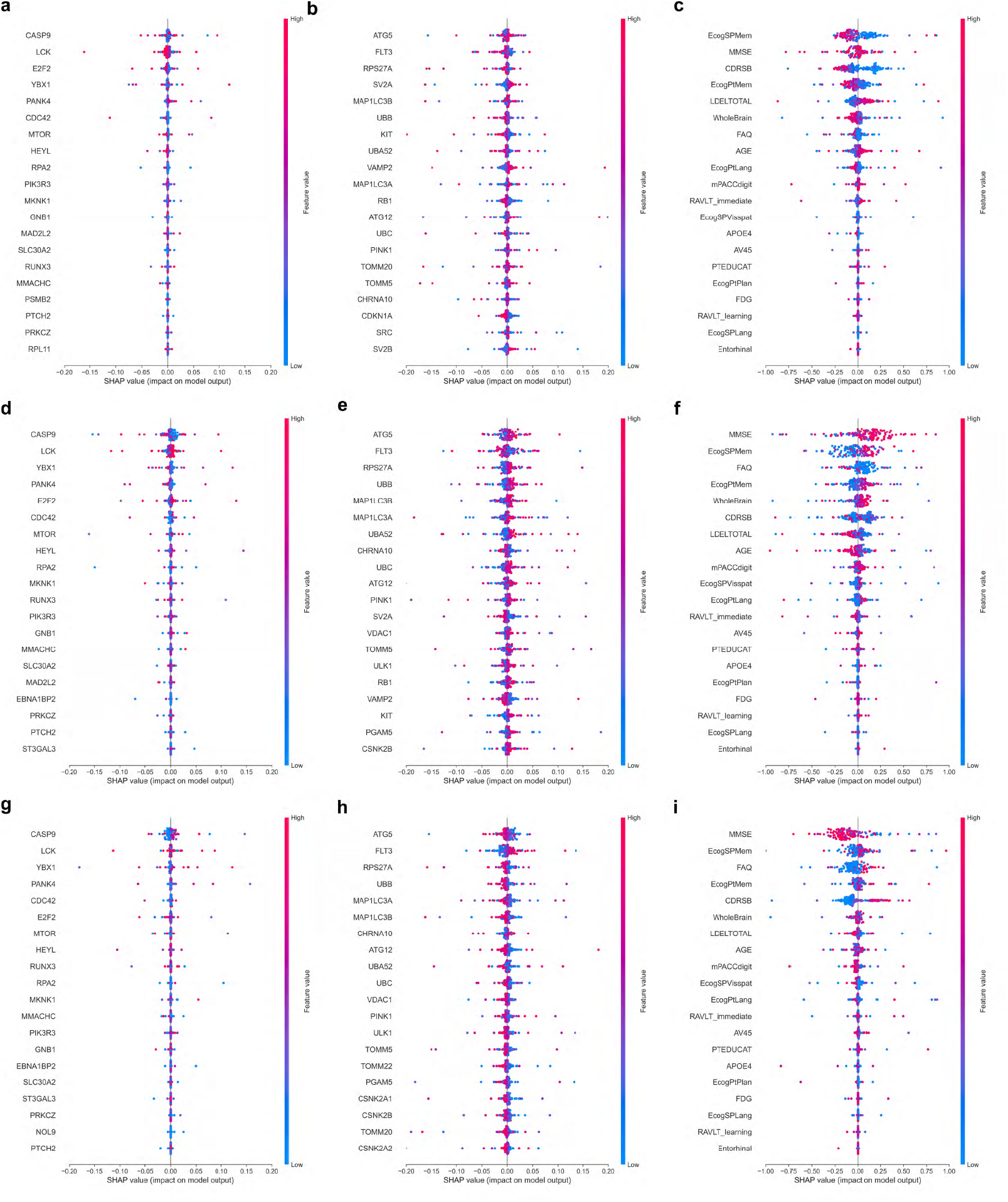
SHAP summary beeswarm plots. SHAP scores colored by feature values are presented for each feature and prediction class. Plots show the impact of top 20 **(a, d, g)** aggregated SNPs features, **(b, e, h)** microarray gene expression data, and **(c, f, i)** clinical data for **(a, b, c)** CN, **(d, e, f)** MCI and **(g, h, i)** AD prediction, respectively.

### SNPs

The SHAP beeswarm plots for SNPs are depicted in Figures 1a for CN prediction, 1d for MCI prediction, and 1g for AD prediction. SNPs found in genes such as CASP9, LCK, and SDC3 have been prioritized by the SHAP scores, showing their influence on model behavior. We performed a network analysis with STRING (Search Tool for Interacting Genes/Proteins) using the selected genes to identify enriched biological pathways, as described in the Methodology section 4.6. The resulting network contains 40 edges (protein-protein interaction (PPI) enrichment p-value 0.0157), which suggests that the genes are biologically connected. More than three-fourths of the interactions have been experimentally validated or extracted from curated databases. Notably enriched pathways include cellular response to stress (network strength (NS) of 0.92 with false discovery rate (FDR) of 0.00018) and CD28 costimulation (NS 1.74 with FDR 0.00055). CD28 is expressed by T-cells and is essential for T-cell proliferation and cytokine production.

### Gene expression

The SHAP beeswarm plots for gene expression are depicted in Figures 1b for CN prediction, 1e for MCI prediction, and 1h for AD prediction. The expression of CCNE1, ATG5, MAP1LC3B, RB1, UBC, TOMM20, and PINK1 are marked as significant for model prediction with SHAP. We examined the protein-protein interaction network using STRING to identify enriched pathways among the selected genes. The network contains 164 edges (PPI enrichment p-value < 1.0 × 10^−16^), with the vast majority being experimentally validated and/or extracted from curated databases, indicating biological relevance. Enriched pathways include aggrephagy (NS 2.12, FDR 0.0082), mitophagy (NS 1.99, FDR 0.0119), autophagosome assembly (NS 1.52, FDR 9.02 *×* 10^−5^) and selective autophagy (NS 1.69, FDR 0.0027). Further, pathways related to DNA damage response (NS 1.78, FDR 7.36 *×* 10^−7^) and nucleotide repair (NS 1.83, FDR 9.27 *×* 10^−5^) have been enriched.

### Clinical features

The SHAP beeswarm plots for clinical features are depicted in Figures 1c for CN prediction, 1f for MCI prediction, and 1i for AD prediction. We observed that the clinical features, such as MMSE, Ecog, FAQ, RAVLT immediate recall, RAVLT learning and CDRSB scores are highly impactful in prediction of AD. Other clinical features with significant influence on model prediction include whole brain volume, presence of APOE *ϵ*4 allele, age, and uptake levels of FDG and AV45.

### Intermediate layer activation

Nodes with significant differences in activation across CN, MCI, and AD inputs represent pathways through which genotyping and gene expression inputs impact c-Triadem predictions. Therefore, we examined node activation in the intermediate layers of the subnetworks. We observed differences in activation at all three hierarchy levels in our model. Figure 2 depicts the kernel density estimation curves for pathway nodes in the intermediate layers of the subnetworks with significant differences in activation. At the lowest level, we observed differences in activation among the nodes representing PLC*γ*1 (phospholipase-c gamma 1) signaling, neutrophil degranulation, and TRKA (tropomyosin receptor kinase A) activation by NGF (nerve growth factor). Differentially activated pathways at the second level include DNA double-stranded break response, formation of apoptosomes, and caspase activation via death receptors. Third-level pathways include cytochrome-c mediated apoptotic response, caspase activation, and signaling by NOTCH and FLT3.

**Fig 2.**
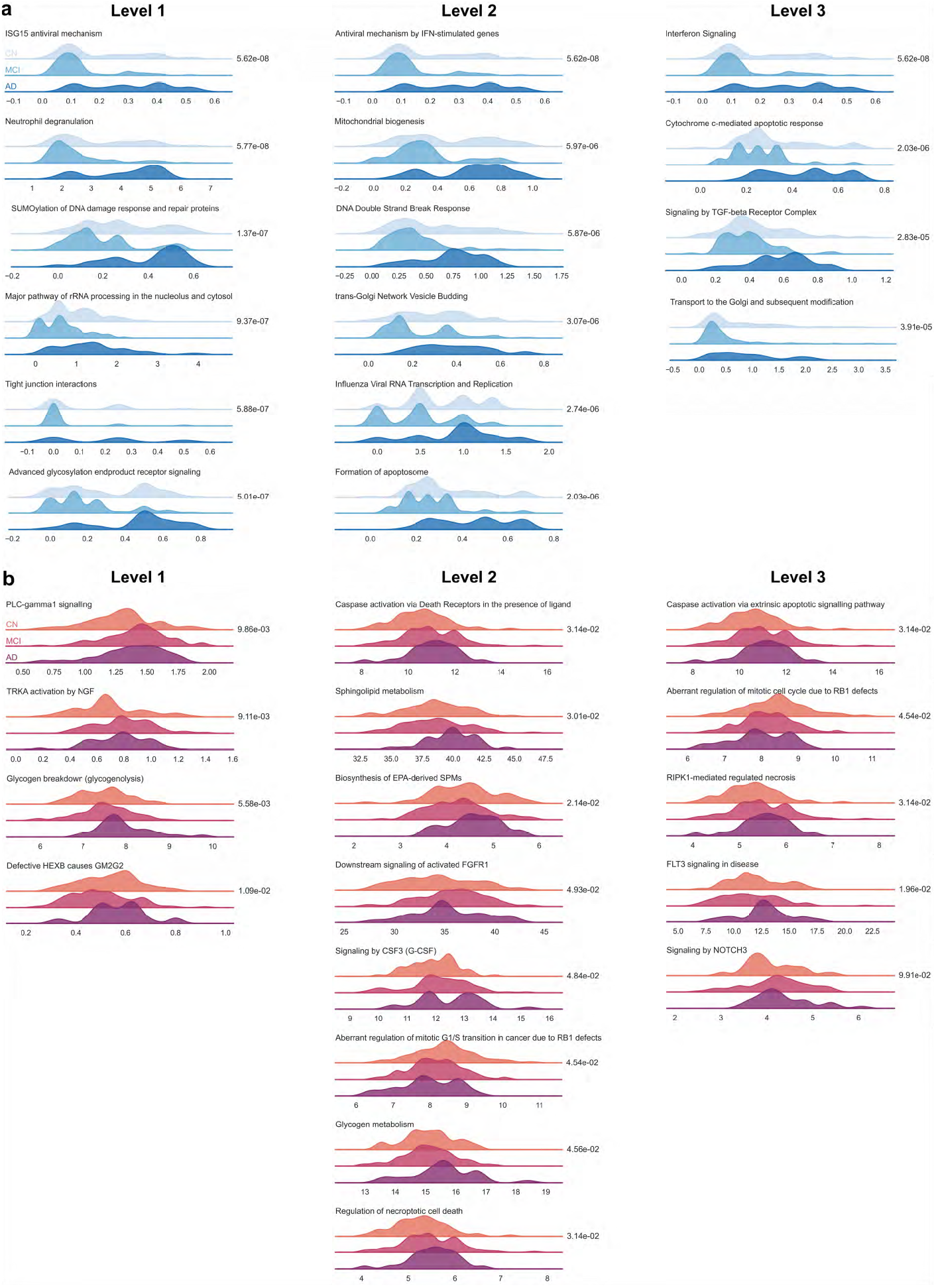
Kernel density estimation curves for intermediate layers. Kernel density estimation curves are presented for pathways with significant differences in activation (established by the Kruskal-Wallis test) at all three levels of hierarchy for (a) SNPs and (b) gene expression data. The P-value for each pathway is reported adjacent to the graph.

## Discussion

In this study, we have described our model c-Triadem, a constrained multimodal deep neural network multiclassifier that accurately predicts the patient’s disease status as either CN, MCI, or AD on the basis of their genotyping, gene expression, and clinical data. The input layers contain nodes encoding genes and their respective biological pathways, and edges are constrained to reflect known relationships among the genes and pathways. The pathway information was taken from the Reactome pathway database, which systematically associates proteins with their functional role in biological pathways, and connects pathways hierarchically. We have shown that our model has superior performance compared to previous deep neural networks trained with the ADNI dataset.

SHAP scores enable the identification of genetic and clinical features which explain the model predictions. From the STRING interaction networks, it is observed that the genes prioritized by SHAP (such as CASP9, LCK, SDC3, CCNE1, ATG5, UBC, and TOMM20) are associated with each other in AD-related biological pathways. The gene CASP9 (caspase-9) is a cysteine-aspartic protease that serves as an initiator for apoptosis. CASP9 is well-studied in neurodegenerative diseases, with increasing levels of CASP9 implicated in AD brains as well as platelet-rich plasma. [33–35] LCK (lymphocyte-specific protein tyrosine kinase) is located in a well-known linkage region [36] and has been named a risk factor for AD. [37] Mouse studies suggest that LCK inhibition alters spatial learning and impairs long-term memory. [38] SDC3 (Syndecan-3, a proteoglycan) expression in PBMC has been correlated with A*β* and has been suggested as a potential biomarker for the diagnosis of early AD. [39] Several studies have identified dysfunctional mitophagy as an early marker of Alzheimer’s disease, driving amyloid-*β* aggregation and neurofibrillary tangle formation. [40] Mitophagy and aggrephagy refer to the autophagic clearance of damaged mitochondria and protein aggregates respectively. Reduced levels of mitophagy markers ATG5 and Parkin have been demonstrated in MCI and AD patients. [41] Given that hypometabolism caused by impaired energy metabolism is one of the earliest possible markers of AD, genes related to mitophagy may serve as potential biomarkers for AD diagnosis in the early stages. Additionally, SHAP-selected clinical features (RAVLT scores, specifically immediate recall and percent forgetting) have a strong association with brain structural atrophy. [42]

The constrained model also allows the identification of biological pathways which emphasize the differences among the three diagnostic groups. Moreover, the selected pathways with significant differences in intermediate node activation (such as PLC*γ*1 signaling, TRKA activation by NGF, DNA double-stranded break response, and caspase activation) reinforce SHAP findings. For instance, SHAP-selected CASP-9 works through the caspase activation pathway identified by intermediate node activation. The findings are also supported by previous studies. It is known that PLC*γ*1 activation is reduced in PBMC samples from AD patients. [43] Deficiencies in the TRKA/NGF axis are implicated in the depletion of cholinergic neurons and cognitive decline in AD. [44]

Recent studies indicate that the development of AD is associated with systemic changes in the neuronal environment reflected in other parts of the body. Studies in peripheral blood mononuclear cells (PBMCs) from amnestic MCI and AD patients show differential expression of senescence markers, such as cell cycle blockade (p16 and p53) and DNA damage response (*γ*H2AX). [18] Garfias et al. [45] have reported significantly higher levels of activated lymphocytes in AD patients. Moreover, a gene expression analysis of PBMC samples in the AddNeuroMed cohort [46] identified DEGs (differentially expressed genes) significantly enriched in pathways related to T-cell and neutrophil activation in immune response, lymphocyte differentiation, protein serine/threonine kinase activity, GTPase and DNA transcription factor binding. [47] In our study, we have identified potential genetic markers of amyloidosis, immune activation, DNA damage response, and dysfunctional mitophagy in blood-derived genetic data. Further, the information from enriched pathways provides mechanistic insights into AD pathogenesis.

While our model has been validated through network analysis and consistency with literature reports, we recognize that multimodal data from additional cohorts would corroborate our findings. As more data become available, we hope to refine our model and establish its validity on other independently curated datasets. We also intend to incorporate non-coding SNP data and other data modalities to improve performance, as non-coding SNPs may affect gene regulation and expression indirectly, thus revealing more mechanistic insights into AD pathology. Our model uses explainable AI to probe multi-omic data for genes and clinical features associated with AD pathogenesis and provides further transparency by exposing the biological pathways which influence its predictions. As a result, we firmly believe that c-Triadem holds the potential to facilitate precise and early diagnosis of AD, as well as other forms of dementia.

## Materials and methods

### Dataset

#### Alzheimer’s Disease Neuroimaging Initiative

Data used in the preparation of this article were obtained from the Alzheimer’s Disease Neuroimaging Initiative (ADNI) database. ADNI was launched in 2003 as a public-private partnership, led by Principal Investigator Michael W. Weiner, MD. The primary goal of ADNI has been to test whether serial magnetic resonance imaging (MRI), positron emission tomography (PET), other biological markers, and clinical and neuropsychological assessment can be combined to measure the progression of mild cognitive impairment (MCI) and early Alzheimer’s disease (AD). For up-to-date information, see https://adni.loni.usc.edu/. In addition to MRI and PET neuroimaging of patients at regular intervals, ADNI has collected and analyzed whole blood samples for genotyping and gene expression analysis. Blood gene expression profiling was conducted using Affymetrix Human Genome U219 Array for 744 samples in the ADNI2/ADNI-GO (Grand Opportunity) phase. Blood genotyping and gene expression methods for ADNI have been described in detail by Saykin et al. [29] The ADNI studies were approved by institutional review boards of all participating institutions, and written informed consent was obtained from all participants or authorized representatives. [29]. The study protocol obtained approval from the human research committees at each involved institution, and informed consent was provided by all participants or their legal guardian(s)/legally authorized representatives. The research procedures strictly adhered to applicable guidelines and regulations [52, 53]. Table 4 presents a summary of the genotyping data provided by ADNI.

**Table 4.** ADNI genotyping data summary. ADNI-GO: Alzheimer’s Disease Neuroimaging Initiative Grand Opportunity.

| Phase | Platform | Variants | Participants | Genome assembly | DbSNP build |
| --- | --- | --- | --- | --- | --- |
| ADNI1 | Illumina Human 610-Quad BeadChip | 620901 SNP and CNV markers | 757 | hg18 | 129 |
| ADNIGO/ADNI2 | Illumina Human OmniExpress BeadChip | 730525 SNP and CNV markers | 793 | hg18 | 129 |
| ADNI3 | Illumina Omni 2.5M (WGS Platform) | 759993 SNP and CNV markers | 327 | hg38 | 155 |

We selected sample data from 626 participants that had both SNP data and gene expression data for training, testing, and validation in the deep learning model. Out of 626 participants, 212 were CN, 317 participants had MCI and 97 were diagnosed with dementia due to AD. The criteria for AD and MCI diagnosis in ADNI are detailed by Petersen. [48]

#### Genotype data

Genotyping data were downloaded in PLINK binary format, consisting of `.bed, .bim` and `.fam` files. The `.bed` file is the primary representation of genotype calls of biallelic variants. The `.bim` file accompanies the `.bed` file and provides extended variant information, i.e., SNP IDs, base-pair coordinates, and allele information. The `.fam` file provides sample information, including parent roster IDs and phenotype. We utilized the `.bim` file to identify the presence of variants and encoded SNPs using additive representation (i.e., 0 = homozygous dominant, 1 = heterozygous, 2 = homozygous recessive). We used the dbSNP ID as the unique identifier for SNPs. We then mapped SNPs that occur in coding sequences (CDS) to their corresponding gene loci. Input values represent the aggregate additive value of all SNPs mapped to the coding sequences of individual genes. The SNP data along with gene expression data represent two of the inputs for the model.

#### Clinical data

Clinical data include demographic information, scores from neuropsychological tests, brain volume measurements, and levels of clinical biomarkers of AD, such as A*β*, tau protein, and FDG uptake. Patient demographic information includes age, gender, ethnicity, racial category, marital status, and years of education. The demographic data were encoded as categorical variables before model training. Genotype information (ie. presence of APOE *ϵ*4) is also included. Brain functioning as measured by fluorodeoxyglucose (FDG), PIB (Pittsburgh compound B), and amyloid detection ligand (AV45) uptake with PET are reported. Cerebrospinal fluid (CSF) biomarker levels of A*β*, tau, and P-tau protein have also been recorded. Additionally, the clinical data reports patient scores from a battery of neuropsychological tests. A Functional Activities Questionnaire (FAQ) assesses the patient’s level of independence to perform daily tasks. Everyday cognitive evaluations (Ecog) of the patient’s ability to carry out everyday tasks are reported by the patient (self) and a study partner. Reported scores for Mini-Mental State Exam (MMSE), Montreal Cognitive Assessment (MOCA), Rey’s Auditory Verbal Learning Test (RAVLT), Alzheimer’s Disease Assessment Scale (ADAS), Modified Preclinical Alzheimer Cognitive Composite (mPACC), Digit Span memory test (DIGITSCOR), Trail Making test (TRABSCOR) and Logical Memory Delayed Recall Total Number of Story Units Recalled (LDELTOTAL) are used to estimate the severity and progression of cognitive and memory impairment. MRI measurements of hippocampal, intracranial, mid-temporal, fusiform, ventricle, entorhinal, and whole brain volume are also reported.

In the ADNI clinical data, PIB levels, Ecog scores, A*β* levels, tau protein levels, and DIGITSCOR were missing for a substantial proportion of patients. Therefore, we utilized k-nearest neighbors (kNN) imputation for handling missing data. kNN imputation selects k subjects that are similar to the subject with missing values and is preferred for its ability to handle continuous, categorical, and discrete data in our dataset. Moreover, kNN imputation is shown to improve multiclass prediction of disease in ADNI. [32] We performed imputation in R v4.2.1 using DMwR (data mining with R) package v0.0.2, on clinical features with less than 60% missing data. We set k = 5 as the minimum number of neighbors from which the missing values could be inferred. Imputation was performed on the training dataset and then applied to the test and validation datasets. Only clinical features measured at the time of diagnosis were retained, ie., baseline features were removed. The target variable (i.e., diagnosis of CN, MCI, or AD) was not included in the procedure.

### The proposed c-Triadem model

The proposed c-Triadem is a constrained artificial neural network multiclass classifier which aims to accurately predict the patient’s status as CN, MCI, or AD using their genotyping, gene expression, and clinical data. The data pre-processing, model development, and interpretation steps involved in c-Triadem are depicted in the block diagram of Figure 3. In particular, in c-Triadem, the genotype and gene expression inputs consist of nodes that represent genes and are fed into constrained subnetworks. The nodes and edges of the hidden layers in the two subnetworks (for genotype and gene expression) represent Reactome pathways and their biological relationships, respectively. We chose to incorporate Reactome into our deep learning model due to its hierarchical organization and a data model which makes pathways computationally accessible. [49] Reactome data was provided by Elmarakeby et al. [50] in their repository (https://zenodo.org/record/5163213#.Y7wZgNVBxPY). By representing the genes, pathways, and connections among them as nodes and edges in the subnetworks, we can better understand the biological connections which are important for the prediction. Thus, the model’s interpretability is enhanced compared to a dense network with similar architecture.

**Fig 3.**
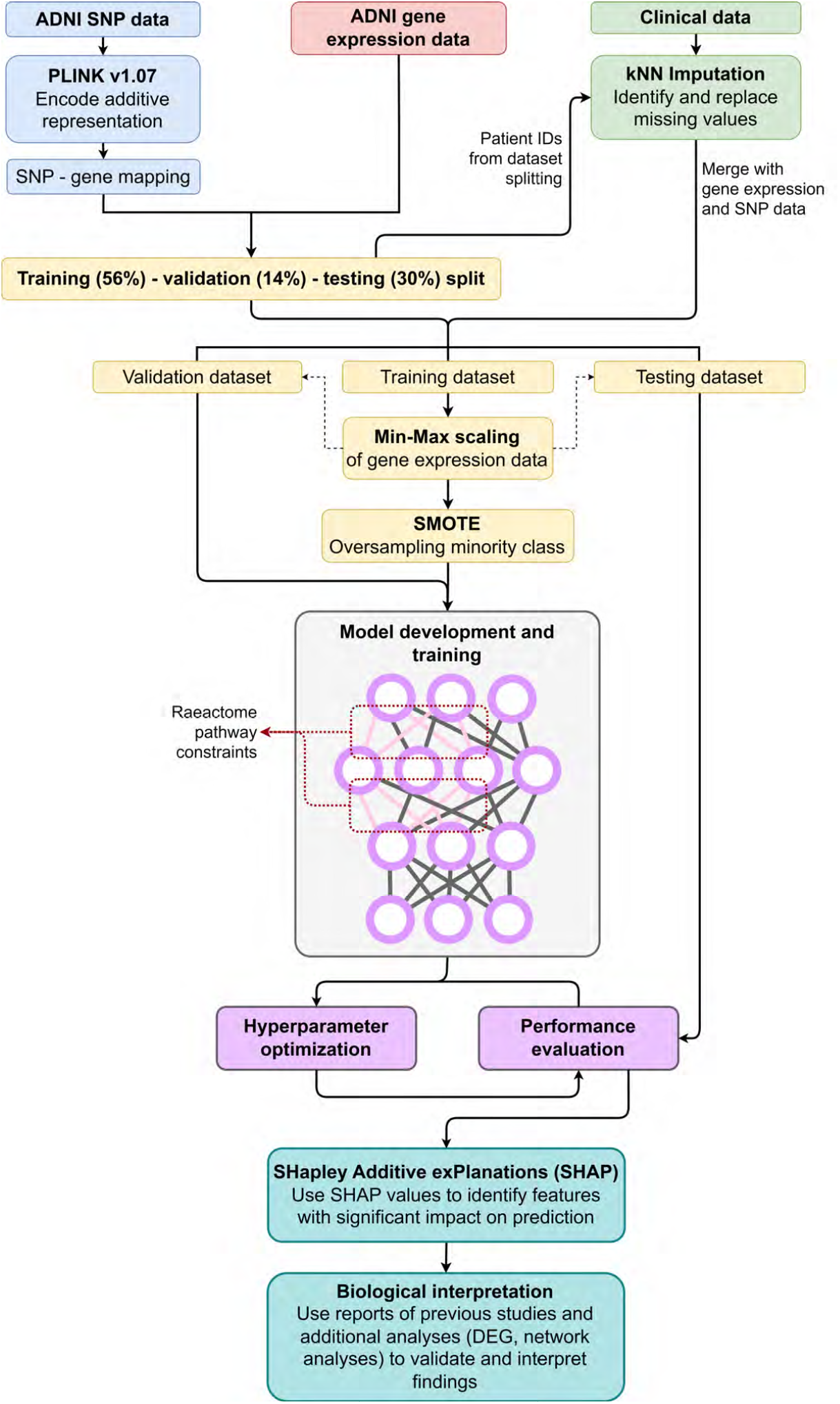
Methodology block diagram. An overview of data preprocessing, model development, and interpretation is presented in the flow diagram.

Each subnetwork consists of one input layer with 10,151 nodes representing the genes and three non-trainable hidden layers with nodes representing pathways. Due to the constraints on the edges connecting the genes and pathways, the sparsely connected subnetwork has 32,842 parameters. The constraints are encoded as a binary weights matrix which sets all non-existent connections among the genes and pathways to zero.

In addition to the genotyping and gene expression input, a third input of clinical data is provided. The clinical data with 45 nodes are concatenated along with the output of the two subnetworks and passed through a batch normalization layer, followed by two hidden layers. Kernel regularization is applied to both hidden layers. Bayesian hyperparameter optimization was used to configure the hidden layer sizes, initial learning rate, step interval for learning rate schedule, choice of activation function, and kernel regularization of the hidden layers, by monitoring the validation accuracy over 80 epochs. A random state was set beforehand to ensure consistent results. The first hidden layer is configured with 19 nodes and linear activation followed by another hidden layer with 8 nodes and Rectified Linear Units (ReLU) activation. A dropout layer (rate = 0.597) is included in between the hidden layers. Overall, our network contains 44,239 parameters and 15 layers. Excluding the non-trainable parameters in the subnetworks, we have 11,397 trainable parameters for three types of input representing 20,347 features, whereas an unconstrained network with similar architecture contains over 37 million parameters.

### Model training

We empirically observed a four-fold reduction in training time for our model (training time of 0 minutes and 18.905 seconds) compared to the unconstrained model (1 minute and 18.862 seconds) on an Intel Core i5 8th Gen CPU (central processing unit) with a clock speed of 1.6-1.8 GHz. Our model was compiled with an adaptive learning rate initially set at 0.008 with exponential decay occurring every 17 steps at a rate of 0.96.

Model training was performed with the Adam optimizer to reduce categorical cross-entropy loss. We used Python v3.8 with the Functional API (Application Programming Interface) of keras v2.4.3 to design and train c-Triadem on genotyping, gene expression, and clinical data from ADNI participants. The dataset comprises 212 CN, 317 MCI, and 97 AD samples. We applied a train/test/validation split of 56-30-14%. We used target-based stratification during the test-train split followed by Synthetic Minority Over-sampling Technique (SMOTE) to address the imbalance in the datasets. Min-max scaling was performed on the training data and transferred to the validation and test datasets. The target was one-hot encoded prior to training.

Along with kernel regularization and dropout, early stopping callbacks on validation loss were used to prevent overfitting. We used the default batch size of 32 and set the maximum number of epochs at 120. The output is converted to respective CN, MCI, and AD probabilities by softmax, with the higher probability used for the classification of disease status. A representation of the model architecture is provided in Figure 4.

**Fig 4.**
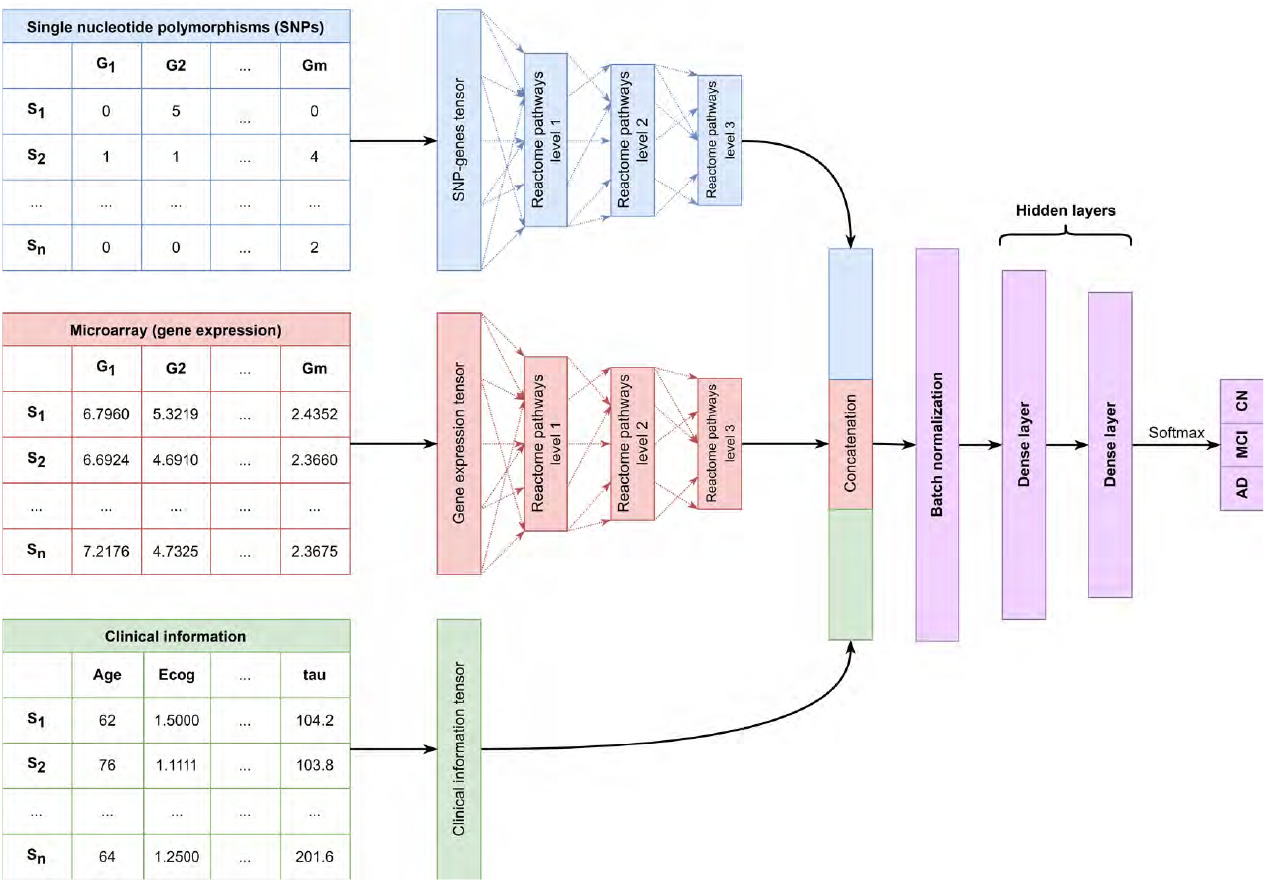
c-Triadem model architecture. *S*_*n*_ represents the nth sample and *G*_*n*_ represents the nth gene. Dummy values are provided for input data types.

### Model performance evaluation

We evaluated the performance of our model on the validation and test datasets. We used the area under the receiver operating characteristic (AUC) curve, accuracy, precision, recall, and the F1 score as performance metrics, and their formulae are listed below. We defined true positives (TP), true negatives (TN), false positives (FP), and false negatives (FN) for each predicted class (CN, MCI, and AD). For instance, for the AD class, TP is the number of AD samples predicted correctly. TN is the number of non-AD samples predicted as MCI or CN. FP is the number of MCI and CN samples predicted as AD. FN is the number of AD samples predicted as MCI or CN.

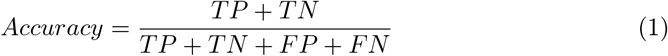

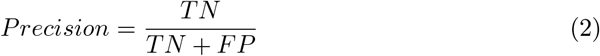

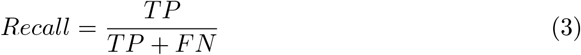

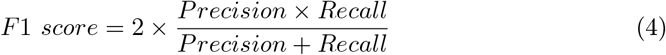

### Model interpretation and identification of potential biomarkers

Model interpretation is essential to gain user trust and overcome the ‘black box’ reputation of deep learning models. Lundberg and Lee [27] proposed SHAP values as a unified measure of feature importance, computed using game theory. To calculate SHAP values, each individual feature’s contribution to the predicted value is estimated by comparing predictions over different combinations of features. The SHAP value for a feature is the average of all the marginal contributions to predictions from all possible feature combinations. To evaluate the importance of specific genes and clinical data, we computed SHAP values on all features using the shap package v0.41.0%.

### Interaction network analysis

In order to gain mechanistic insights and validate the reliability of our model, we thoroughly investigated the interaction network among genes that were prioritized by SHAP. Specifically, we extracted the SHAP-prioritized genes from the top 20 features for each group (CN,MCI, and AD). To explore the functional associations between these genes, we employed STRING, a resource that leverages diverse types of evidence, including experimental data, to identify such connections. By examining the interaction network within STRING [51], we aimed to elucidate the intricate relationships and potential collaborative roles among these genes. Moreover, we also conducted an analysis to identify enriched pathways for the selected genes. This allowed us to gain further insights into the biological processes and molecular mechanisms underlying the observed gene interactions. By comprehensively studying the gene interaction network and identifying enriched pathways, we aimed to reinforce the validity and biological relevance of our model’s predictions.

### Intermediate layer activation

To investigate and compare the activation values of intermediate layers in our model, we randomly selected inputs from each prediction class, namely cognitively normal (CN), mild cognitive impairment (MCI), and Alzheimer’s disease (AD). In this context, activation refers to the signed outcome of a node based on its inputs. To determine the significance of differences in activation values, we conducted the Kruskal-Wallis statistical test. This analysis allowed us to assess the variations in activations across the different prediction classes in a rigorous and quantitative manner.

### Statistical analysis

Significant distinctions in clinical features among cognitively normal (CN), mild cognitive impairment (MCI), and Alzheimer’s disease (AD) subjects were examined through rigorous statistical analyses. The age of onset and years of education were subjected to an analysis of variance (ANOVA) test to assess their significance. Similarly, the proportion of male and female participants, as well as the presence of the APOE *ϵ*4 allele, were evaluated using the Chi-square contingency test. For the remaining clinical variables, the non-parametric Kruskal-Wallis test was employed. It is important to note that all statistical analyses were conducted with a significance level of 95

## Data Availability

All data produced in the present work are contained in the manuscript.

## Acknowledgments

Data used in the preparation of this article were obtained from the Alzheimer’s Disease Neuroimaging Initiative (ADNI) database (adni.loni.usc.edu). As such, the investigators within the ADNI contributed to the design and implementation of ADNI and/or provided data but did not participate in the analysis or writing of this report. A complete listing of ADNI investigators can be found at http://adni.loni.usc.edu/wp-content/uploads/how_to_apply/ADNI_Acknowledgement_List.pdf. Reactome data were downloaded from the repository provided by Elmarakeby et al. (2021): https://zenodo.org/record/5163213#.Y7wZgNVBxPY.

## Author contributions statement

SJ: Data curation, Formal analysis, Investigation, Methodology, Software, Visualization, Writing – original draft preparation, Writing – Review and editing. AS: Conceptualization, Data curation, Funding acquisition, Software, Project administration, Supervision, Validation, Writing – Review and Editing. FA edited and reviewed the second manuscript. All authors reviewed the manuscript.

## Data availability statement

The constrained model is made available on GitHub at https://github.com/Sherlyn-J/KU-BMED2/. The dataset analysed in this study is publicly available in the Alzheimer’s Disease Neuroimaging Initiative (ADNI) repository https://adni.loni.usc.edu/. (Accession Number:sa000002).

